# Improving health care worker’s compliance with traceability by recording the nursing process at the point of care using a personal digital assistant with a barcode

**DOI:** 10.1101/2020.01.14.20017434

**Authors:** Olga Florea, Jean Charles Dufour, Chloe Magnin, Philippe Brouqui, Sophia Boudjema

**Affiliations:** Aix Marseille Université, IRD, MEPHI, IHU-Méditerranée Infection, Marseille, France; AP-HM, IHU-Méditerranée Infection, Marseille, France; Aix Marseille Université, AP-HM, INSERM, IRD, SESSTIM, Hop Timone, BioSTIC, Marseille, France

**Keywords:** electronic records, nursing records, personal digital assistant, adverse event, hospital-acquired infection, blood catheter, urinary catheter, traceability

## Abstract

**Background:** Adverse events are serious and frequent complications most often linked to the quality of nursing care.

**Purpose:** We evaluated the compliance to traceability of nursing care at bedside using the Patient Smart Reader®, a personal digital assistant with a barcode.

**Methods:** We compared paper record forms, specific computer software in the hospital information system and the Patient Smart Reader®.

**Results:** The Patient Smart Reader enhanced the recording of 90% of nursing care surveyed. Regarding the insertion of blood catheters, compliance rates increased from 44.19 % to 100%, and blood catheter monitoring has risen from 29.64% to 80.74%. Urinary catheter monitoring and insertion recording increased from 10.23% to 55.43% and from 16.67% to 100 % respectively.

**Conclusions:** Providing to caregivers a nursing record system using barcoded implements at the point of care in real time significantly improved traceability of the nursing care.

## INTRODUCTION

Adverse event (AE) is any unexpected events that follow any act or action performed or prescribed by a health professional. Adverse events are widespread and have a significant cost, it was estimated that the total costs of preventable AEs in the USA was in between $17.1 billion and $29 billion annually ^1^. The two most frequent classes of AE, postoperative infections and pressure ulcers, accounted for the largest annual costs (6.5 billion USD) ^2^. The prevalence of AE can vary from 7 to 40 %, but it is important to note that they could be avoided in more than half of cases and that 27.6% are related to negligence and 76.8% to inappropriate nursing care ^2 3^. In intensive care units (ICU), AE is associated with a more extended hospital stay ^4^. In Canadian hospitals, a study reported 20.8% of AE-related deaths and the authors estimated that the death rate due to AE in Canada as a whole would be > 38,000 annually ^5^. The expected goal in caring for patients is to provide low mortality, low morbidity, and a low readmission ratio after 30 days, with a better quality of life. The Institute of Medicine suggests that improving data collection and analysis of direct patient care would enhance patient safety ^6^. According to the “knowing how to prevent” adage, traceability of care is likely to be a significant component of the surveillance and prevention of AE, suggesting that better traceability should lower AE and their consequences. The use of the ‘Check List’ in anesthesia demonstrated a significant reduction in mortality ^7^. It is suggested that as many as 70% of adverse events could be avoided if the right information about the right patient is available at the right time and the health information exchange makes this possible. In agreement with the recommendations of most national authorities, the care provided by healthcare workers (HCW) is currently registered in the patient’s hospital Electronic Health Record (EHR). The quality of nursing care is related to the execution of the nursing process, which should be adequately documented ^8^. The transfer from paper-based to electronic documentation is commonly available worldwide nowadays. Whether computerized or not, the act is, however, most often recorded manually and outside of the location of the nursing care (the patient’s room), giving place to forgetfulness and inaccuracy. While many studies in the world evaluate the impact of the EHR on nursing care quality, few evaluate specifically the pertinence of a bedside care recorder for the quality of traceability. Comparison of PDA records at the bedside with paper formats suggests that PDA used at bedside is reliable, allowing fewers errors, easy to use, and advocated by interviewers ^9 10^. To improve the quality of nursing care records, the nursing care act should be made in real time, meaning that the bedside solution would be promoted. Some studies report point of care nursing record evaluations ^11, 12^ but quantitative analysis for compliance of traceability of nursing care records has not yet been reported in prospective comparative case-control protocol.

This researcher developed a personal digital assistant solution with a barcode to trace nursing care at the patient’s bedside in real time that was named the Patient’s Smart Reader® (PSR). The study aim to evaluate the impact of such tool on compliance of HCW’s with recording nursing care in the care unit by comparing successively different nursing care recording tools. The hypothesis was that recording care at bedside in real time, will improve the compliance to tracaeability of nursing care.

## MATERIALS AND METHODS

The study was conducted in the infectious and tropical diseases medical unit of the university hospital in Marseille, France, which consisted of 17 beds. For purposes of our research, we decided to divide the unit into two distinct parts, nine beds (for cohort A) and eight beds (for cohort B). All voluntary HCW (doctors, nurses, nursing assistants, and housekeeping personnel) of the unit were included in the study.

### Recorder systems

When an HCW provides nursing care to the patient, he/she must track it on the nursing care record. The validity of this information has to be certified with the time and date of the nursing care and with the signature of the HCW. We compared different traceability systems that were used in patient medical records. The oldest one was a dedicated paper sheet form. This sheet of paper was placed into the paper-based patient’s medical record. The second was introduced in 2013 in our institute and was a specific software, Pharma® (version 5.8.70602.1 300) that allows the traceability of medical prescriptions and drugs administered, but also provides some patient’s parameters such as urine strip, temperature and weight. The third, introduced at the beginning of 2016 in our institute, was an EHR (Axigate® version: 5.6.1P9) embedded in the Hospital Information System (HIS). This EHR allows the registration of some other parameters such as peripheral venous catheter insertion and monitoring, urinary catheter insertion and monitoring, inpatient booklet providing, and isolation monitoring. Finally, the last system, introduced in June 2016, was the Patient Smart Reader (PSR), installed in 8 patient rooms (Cohort B).

The PSR system is a Personal Digital Assistant with a bar code scanner allowing for bedside recording of nursing care (https://vimeo.com/205512348). With the PSR, HCW can record, at the patient’s bedside and in real time, nursing care such as urinary or peripheral venous catheter insertion and monitoring, by using barcoded implements, or can directly record various parameters such as body temperature, blood pressure or pain directly onto the PSR. The PSR provides automatic reminders of controls or alerts. For example, recording the insertion of peripheral venous catheter triggers monitoring every 8 hours automatically. The information collected is transmitted to a server that ensures the storage and facilitates the analysis of data. Reports and alerts were generated by the server and displayed on the main screen available in the staff room, as well as on the PSR.

### Data collection

First, through a before/after study, we retrospectively compared the compliance of data reported on single sheets from April 2012 to March 2013 (period 1), to that collected into both Pharma® software and paper sheets from March 2013 to April 2014 (period 2). Then, we compared in a prospective cohort study, from June 2016 to January 2017, the compliance to traceability using our institutional available systems; paper sheets plus Pharma®software and EHR (Cohort A) against the PSR alone (Cohort B). For this purpose, HCW were asked to use only the PSR in the eight equipped rooms to record care.

### Data analysis

We used descriptive statistics, and we described data compliance in terms of frequencies and percentages. Compliance to traceability (as defined by variable recorded / variable to be recorded) of nursing care using different tools (paper sheet versus paper sheet and Pharma software or any kind of recording systems versus PSR) were analyzed. Differences in compliance obtained were defined significant when p<0.05 (Chi-square test). Similarly, relative risk and 95% confidence interval was reported.

### Variables studied

It was decided to monitor the compliance in traceability of 10 variables selected for their public health impact on the safety of care, or their mandatory systematic nature. Some of these acts are mandatory for all patients (welcome inpatient booklet, urine strip, weight once per hospital stay upon patient entrance and body temperature three times a day, and catheter monitoring when appropriate), but blood cultures, blood and urinary catheter insertion and isolation monitoring were prescribed by a medical doctor.

### Inclusion / exclusion criteria

All patients with a length of stay less to 5 days were excluded from the study. The choice of 5 days has been set to allow a sufficient amount of time in the hospital setting in which a catheter can be prescribed and other nursing care to be needed.

### Ethics

Patients were informed of the study upon admission to the unit. They were informed that this did not affect their medical care. The study was approved by our independent ethics committee under N° 2018-011.

## RESULTS

Medical records for 1532 patients were reviewed, and 732 files were excluded from the study because the length of stay was less than five days. Finally, 800 files were considered suitable for analysis of 18,455 opportunities to assess compliance.

Table 1 summarized the compliance to traceability of the 10 variables between paper forms (period 1; 407 files) and paper forms plus Pharma software (period 2; 393 files). The paper forms reported a low compliance to traceability rate; the highest was blood catheter monitoring (59.07%) and temperature (58.67%). The other were : weight (55.28 %), welcome inpatient booklet (46.93%), blood culture (43.66%), blood catheter insertion (32.51%) and isolation monitoring (31%). Urine strip was traced in 13.51% of cases only, and urinary catheter monitoring showed a meager compliance ratio (0.62%). Following the introduction of the Pharma® software, a significant increase of the compliance to tracability of nursing care records was observed for urinary strips which increased from 13.51% to 19.85% (p=0.02), and temperature from 58.67 % to 87.72 % (p<0.0001). Regarding the other variables, no significant changes were observed, except a slight but significant decrease in compliance with blood catheter monitoring (59.07% to 56.31% p=0.014).

**Table 1:**
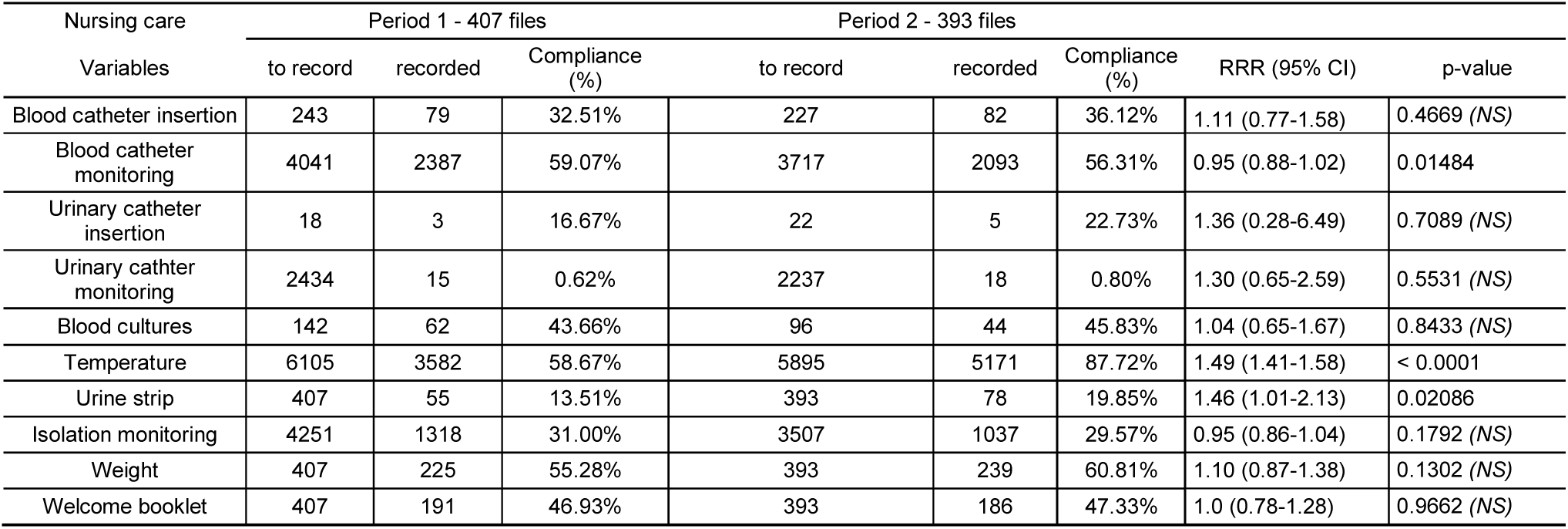
Compliance to traceability of 10 variables between paper forms (period 1) and paper forms plus Pharma software (period 2)

In the prospective cohort study, a total of 341 files were collected, 148 records were excluded from the study according to exclusion criterion, and 193 records were included, 101 in the cohort (A) and 92 in the cohort (B), thus representing 6,073 opportunities submitted for analysis. The use of PSR alone improve significantly the compliance to traceability of all nursing care records but urinary catheter insertion.

Among the essential findings according to relative risk and 95% confidence interval, blood catheter insertion increased from 44.19% to 100% of prescribed insertion, and blood catheter monitoring has risen from 29.64% to 80.74% of catheters to be monitored. Two other items improved significantly, the body temperature from 53.12% to 80.81%, and the isolation monitoring from 26.57% to 43.85% of prescribed isolation (Table 2).

**Table 2:** Compliance to traceability of 10 variables between Any Type of Records* (Cohort A) and PSR (Cohort B)

| Nursing care<br>Variables | Cohort A - 101 files |  |  | Cohort B- 92 files |  |  | RRR (95% CI) | p-value |
| --- | --- | --- | --- | --- | --- | --- | --- | --- |
|  | To traced | Traced | Compliance (%) | To traced | Traced | Compliance (%) |  |  |
| Blood catheter insertion | 43 | 19 | 44.19% | 31 | 31 | 100.00% | 2.26 (1.08-4.71) | < 0.0001 |
| Blood catheter monitoring | 631 | 187 | 29.64% | 244 | 197 | 80.74% | 2.72 (2.12-3.49) | < 0.0001 |
| Urinary catheter insertion | 6 | 1 | 16.67% | 2 | 2 | 100.00% | 6.0 (0.33-107.42) | 0.1071 |
| Urinary catheter monitoring | 88 | 9 | 10.23% | 92 | 51 | 55.43% | 5.42 (2.51-11.66) | < 0.0001 |
| Blood cultures | 63 | 16 | 25.40% | 26 | 20 | 76.92% | 3.02 (1.36-6.74) | < 0.0001 |
| Temperature | 1508 | 801 | 53.12% | 1188 | 960 | 80.81% | 1.52 (1.34-1.71) | < 0.0001 |
| Urine strip | 101 | 12 | 11.88% | 92 | 40 | 43.48% | 3.65 (1.80-7.40) | < 0.0001 |
| Isolation monitoring | 764 | 203 | 26.57% | 358 | 157 | 43.85% | 1.65(1.29-2.1) | < 0.0001 |
| Weight | 101 | 29 | 28.71% | 92 | 69 | 75.00% | 2.61 (1.55-4.38) | < 0.0001 |
| Welcome booklet | 101 | 27 | 26.73% | 92 | 68 | 73.91% | 2.76 (1.63-4.68) | < 0.0001 |

\* Any type of record: variables recorded on any of available Paper forms plus Pharma® software and Electronical Medical Record

Since then, the system has been deployed in the unit, allowing continuous and real-time monitoring of nursing files. Two years after the completion of the study, the average compliance to traceability of nursing care records of some mandatory items such temperature, pulse, blood pressure, and pain was 91.7% (Week 7 February 2019), suggesting that changes in the behavior of healthcare workers have been sustained.

## DISCUSSION

Reviews of PDA usage in health care indicate that they are widely used, functional, and useful for documentation. That PDAs might improve decision making, reduce medical errors, and enhance learning for students and professionals ^13, 14^.

A nursing record system allows the care that has been planned or provided to individual patients by nurses or other caregivers to be recorded. Several studies have already compared paper-based and electronic computerized nursing records, which suggest that electronic systems are preferred ^15 16^. Nursing record systems may be an effective way of influencing nurse practice. However, with the use of these technologies, nurses are expected to change the way they document patient care by shifting from paper forms to electronic systems. While electronic medical records documentation in presence of the patient likely impact nurse patient relation-ship ^17^, the bedside recording with a barcode in our experience allow maintaining this relation. Wang et al. in their study reported that the overall quality of documentation content for the nursing process was not better in the electronic system than in the paper-based system ^18^. Interestingly, in our study, the addition of a new nursing record electronic system such as Pharma® has improved the traceability of body temperature and urine strip recording, but inversely, has significantly decreased the compliance to blood catheter monitoring. However, in our study, the lack of significant improvement in urinary catheter insertion traceability with the PSR compare to any type of records reported in table 2, is likely related to the study power.

To avoid information transmission oversights and improve the compliance to traceability of nursing care, it was suggested that care recording should be done during the care at bedside. Moreover, the fight against cross-transmission of pathogens in hospitals, resulting in hospital acquired infection, explains the choice of a dedicated PDA for each bedroom. Among the weaknesses of our study, the randomization was not feasible, because of health care organization which do not allow a random assignement of patients.

To the best of our knowledge, there is no other study reporting the quantitative evaluation of point of care nursing records. The advantage of recording at the point of care is twofold, being safer from an hygiene point of view (one PDA per room) and avoiding the loss of information between the bedroom and the nearest computer. Moreover, the data collected by the PSR can be directly transferred to the care plan in the patient’s electronic files of the hospital information system. We evaluate ten variables because of their mandatory character, but the PSR contains up to 80 variables and allows for decision-making. For instance, once a nurse has registered the introduction of a peripheral venous catheter, the system asks her if the catheter can be removed each shift (8 hours).

Post operative infection, central and peripheral venous catheter related blood stream infections represent more than half of the cost of all AE ^2^. For this reason we focused our study on variables mostly associated with hospital acquired infection rather than drug prescription or falls, which are the two other most frequent AE reported in the literature. Unfortunately, as the research question was focused on the compliance of tracaeability of nursing care, the adverse events were not recorded.

In conclusion, providing to caregivers a nursing record system using barcoded implements (PSR) at the point of care in real time significantly improved traceability of the nursing care.

## Data Availability

All detailed data are available on request to the corresponding author

## Acknowledgments

The authors thank the healthcare personnel of the medical unit for their commitment and participation in this study.

